# The managerial role and psychosocial factors of job satisfaction: a cross-sectional study among Wittyfit’s users

**DOI:** 10.1101/2022.07.30.22278228

**Authors:** Rémi Colin-Chevalier, Bruno Pereira, Samuel Dewavrin, Thomas Cornet, Marek Żak, Amanda Clare Benson, Frédéric Dutheil

## Abstract

**Background:** Job satisfaction is an emerging indicator for measuring workers’ occupational well-being, however this has been poorly studied in those with managerial roles.

**Objective:** We aimed to explore job satisfaction in workers and to determine and assess psychosocial factors that may influence this relationship.

**Methods:** Data from Wittyfit’s users were collected between January 2018 and February 2020. Volunteers anonymously provided socio-demographic data and responses to questionnaires about their levels of job satisfaction and psychosocial feelings (ambiance, meaning, organization, recognition, values, work-life balance).

**Results:** 10,484 employees (40.9% of women) and 836 managers (33.9% of women), i.e. 11,320 workers with median age of 45 years and seniority of median 10 years of service, were included in the study. Job satisfaction of workers was higher in managers than employees (mean ± SD 68.1 ± 20.4 vs 57.8 ± 24.2, p < .001), as were their feelings about ambiance (71.2 ± 20.9 vs 66.1 ± 24.2), meaning (66.9 ± 21.8 vs 56.1 ± 23.1), organization (55.3 ± 23.6 vs 46.6 ± 24.6), recognition (62.8 ± 23.9 vs 48.3 ± 26.7) and values (66.3 ± 21.7 vs 56.5 ± 23.1) (p < .001). There was no difference in work-life balance (58.1 ± 23.9 vs 59.2 ± 23.4, p = 0.2). All psychosocial factors had an impact on job satisfaction for both managers and employees (p < .001). High job satisfaction was more prevalent in workers who were managers than in employees (84.6 vs 68.8%, p < .001). Even though the managerial position was the most influential factor of job satisfaction (OR = 2.65, 95% CI 2.18 to 3.23, p < .001), other socio-demographic variables such as age (0.87, 0.79 to 0.95, p = 0.002) and seniority (0.71, 0.65 to 0.79, p < .001) also had an influence, although three times less.

**Conclusions:** Managers seem to have higher job satisfaction and psychosocial feelings about their work than employees. Psychosocial factors, many which are modifiable, as well as socio-demographic factors such as age and seniority, may influence job satisfaction among workers.

**Trial registration:** Clinicaltrials.gov: NCT02596737.

## Introduction

Job satisfaction is a widely studied concept, with positive benefits both for workers and companies (1–5). It is a major and increasingly used indicator to assess occupational health of different aspects of work, making it a global importance at the workplace (6). One possible definition of job satisfaction is “a pleasurable or positive emotional state resulting from the appraisal of one’s job or job experiences” (7). In addition to common models used to assess occupational well-being of workers (8–12), job satisfaction is an important attribute in measuring quality of life and stress at work (1,6). Therefore, understanding the predictors of job satisfaction allows for the development of effective strategies to improve occupational well-being and business performance (13). To the best of our knowledge, job satisfaction has been poorly studied in the general population, and nearly no studies assessed the differences in job satisfaction between managers and employees, or they focused only on healthcare professions (14). As a crucial role within a company, it therefore seems necessary to investigate the importance of managerial status on perceived job satisfaction.

There is extensive research around putative psychosocial factors that can influence job satisfaction. Work-related factors such as ambiance (15), meaningfulness at work, values (16), organization, recognition and work-life balance (17) have appeared in the literature as being related to job satisfaction. Despite these by nature being more subjective than other factors like workload or autonomy, they capture the personal feelings of workers. Although they have been assessed individually, to date no study has collectively compared their influence on job satisfaction and according to the job position of the worker.

Thus, the main objective of our study was to explore the influence of the job position (manager or employee) on job satisfaction. The secondary objectives were to assess the other psychosocial factors in relation to the job position and their influence on job satisfaction, as well as the putative influence of socio-demographic factors such as age, seniority and sex (14,18,19).

## Materials and Methods

### Recruitment

This cross-sectional study was conducted among Wittyfit users. Wittyfit is an online platform (20) designed by the Wittyfit company (Paris, France) in partnership with the University Hospital of Clermont-Ferrand to improve the professional well-being of workers (21). Through different questionnaires, volunteers are invited to anonymously report how they feel in several health-related areas when logging into the platform. Wittyfit provides workers with individualized feedback, and managers with a report of their team’s average health status and the issues affecting them. This study was approved by the National Commission for Data Protection and Liberties (CNIL), and the South-East VI ethics committee (clinicaltrials.gov identification number NCT02596737). Due to the COVID-19 pandemic, data collection for this study was limited to the period between January 2018 and February 2020. All workers who completely or partially answered the Wittyfit “job satisfaction” questionnaire at least once during this period were included in the study. Workers who did not fill in their personal information (age, seniority, sex or position in the company) were excluded.

### Outcomes

#### Primary outcome

Job satisfaction of workers was assessed via the Wittyfit “job satisfaction” questionnaire using the visual analogical scale (VAS; score range from 0 to 100). Wittyfit uses VAS to assess health outcomes in the same way as it does to measure stress, for example (22–24). Workers could estimate their job satisfaction as many times as they wanted. For each indicator (job satisfaction and psychosocial feelings), we used the average score to evaluate the feeling of a worker. We considered a worker to be satisfied when their score was higher than 50 and dissatisfied lower than 50.

#### Secondary outcomes

Psychosocial indicators included workers feelings of ambiance, work-life balance (or simply balance), meaning, organization, recognition and values. Ambiance at work assesses the relationship with the manager and colleagues, the quality of the work environment and the social support at work. Work-life balance measures the prioritization of personal and professional activities by the worker. Meaning it evaluated the interest, usefulness, fulfillment and motivation of the worker. Organization assessed the definition of worker’s job description, resources, level of worker responsibility, their confidence in management and colleagues and satisfaction with their salary. Recognition investigated the respect between management and colleagues and the appreciation and acknowledgment of the contributions of the individual to the company. Finally, values explored the balance between corporate and personal values. Similar to job satisfaction, all psychosocial indicators were evaluated using the associated VAS in the questionnaire. Workers could estimate their feeling in each category as many times as they wanted and their average score was retained. Wittyfit’s clients provided the age, seniority, sex and position (manager or employee) of their workers who use Wittyfit. Workers were then divided into two age categories (over or under 45 years old) and two seniority classes (over or under 10 years of service).

### Statistics

Quantitative data (job satisfaction and psychosocial feelings) were expressed in terms of mean ± standard-deviation. In absence of data, assumed to be missing completely at random (25), a mean imputation by client was applied to avoid complete-cases analysis bias (26). Qualitative data (worker’s job satisfaction/dissatisfaction) were expressed using number of participants and associated percentage. First, comparisons between groups (e.g. by sex, by job position) were performed with ANOVA for continuous variables (job satisfaction and psychosocial feelings) and χ^2^ tests for categorical variables (satisfied/dissatisfied workers). Then, associations between endogenous variables (job satisfaction) and exogenous variables (psychosocial and socio-demographic factors) were assessed with linear and logistic mixed regressions respectively. The company to which the worker belongs was considered a random effect. Variance influence factor values of exogenous variables were assessed and revealed no serious multicollinearity (a value below 4 was considered acceptable). Coefficients of linear regression models have been expressed as effect sizes (ES) and confidence intervals (95% CI) using a “refit” method (27) and interpreted according to Funder (28). Coefficients of logistic regression models are expressed as standardized odds ratio (OR) and 95% CI and interpreted according to Chen (29). Subgroup analyses were conducted by job position to compare the primary drivers of satisfaction in workers. Statistical analyses were performed using R (version 4.1.1) (30) in the RStudio (version 1.3.1056) platform. Unless specified, we considered a *P*-value<0.05 as statistically significant.

## Results

### Participants

Of the 41,856 workers using Wittyfit in 2019, 11,320 answered the “job satisfaction” questionnaire at least once during the period. We obtained the data from 6,753 men and 4,567 women, separated as 10,484 employees and 836 managers, with 49.6% of the population under 45 and 56.0% with under 10 years of service, distributed across seventeen French companies (**Fig 1**).

**Fig 1.**
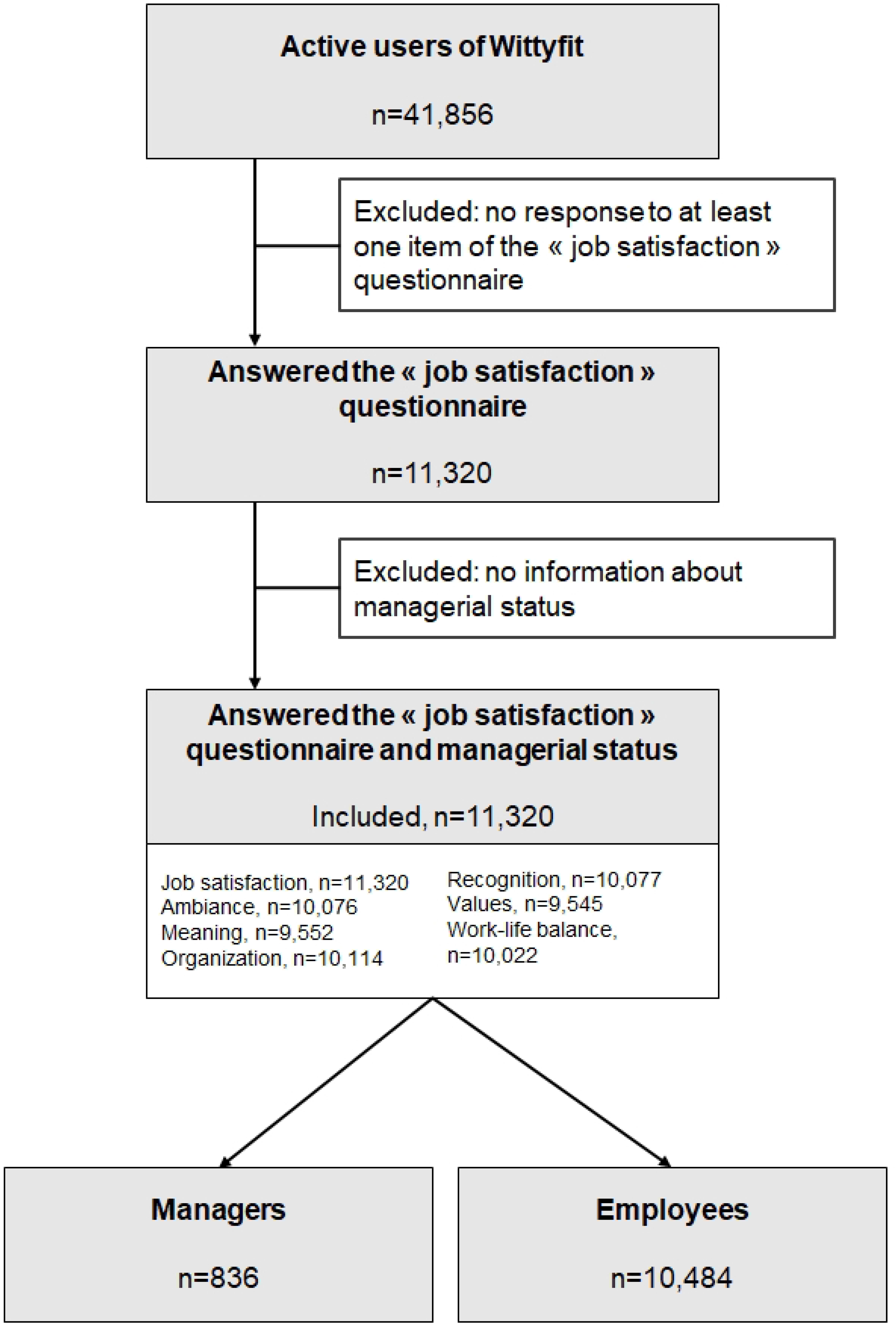
Flow chart of Wittyfit’s users

### Job satisfaction among managers

Managers reported having a higher job satisfaction than employees (68.1 ± 20.4 vs 57.8 ± 24.2, p < .001). Indeed, the position within the company had an influence on job satisfaction (ES = 0.46, 95% CI 0.39 to 0.53, p < .001). Managers also tended to be more satisfied with their jobs, with a higher prevalence of managers highly satisfied with their jobs than employees (84.7 vs 68.8%, p < .001) (**Fig 2**). Managers were twice as likely to be more satisfied than employees (OR = 2.65, 95% CI 2.18 to 3.23, p < .001) (**Fig 3**).

**Fig 2.**
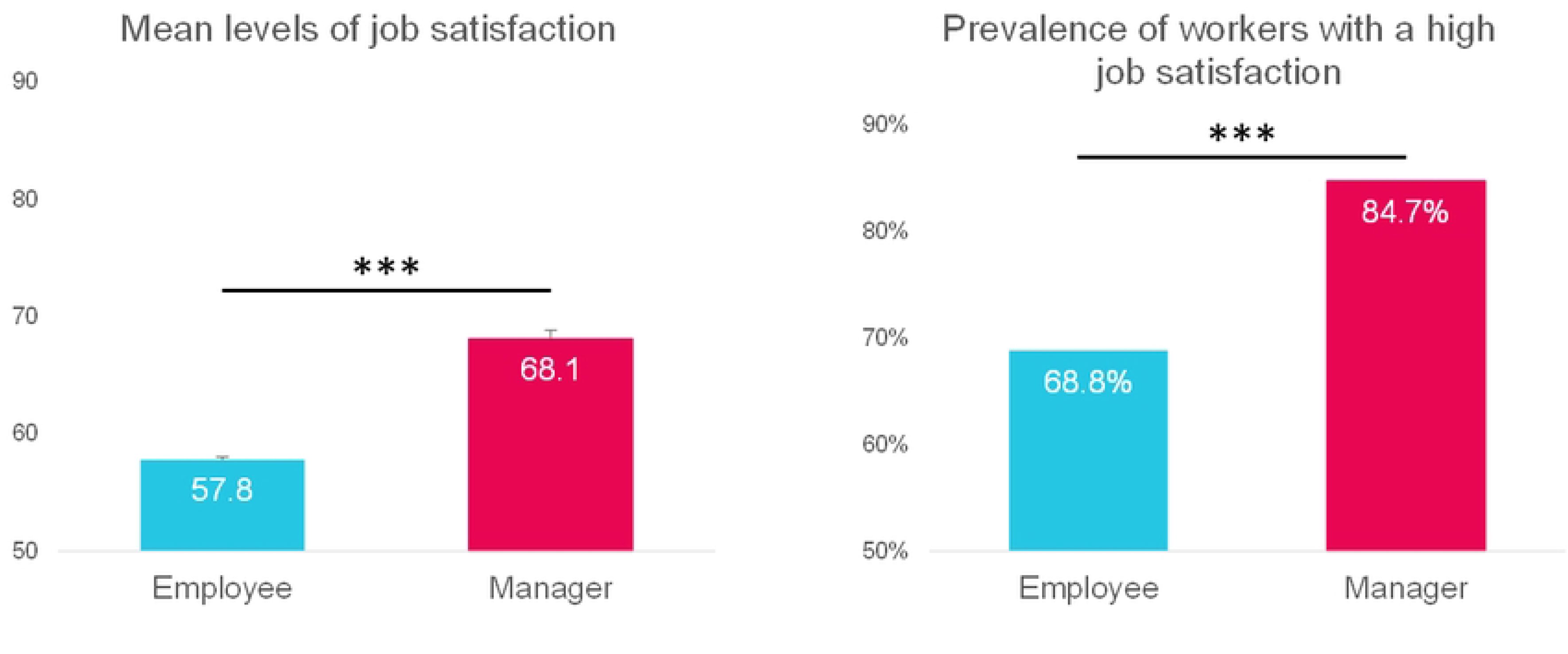
Job satisfaction by job position ‘***’: p < 0.001

**Fig 3.**
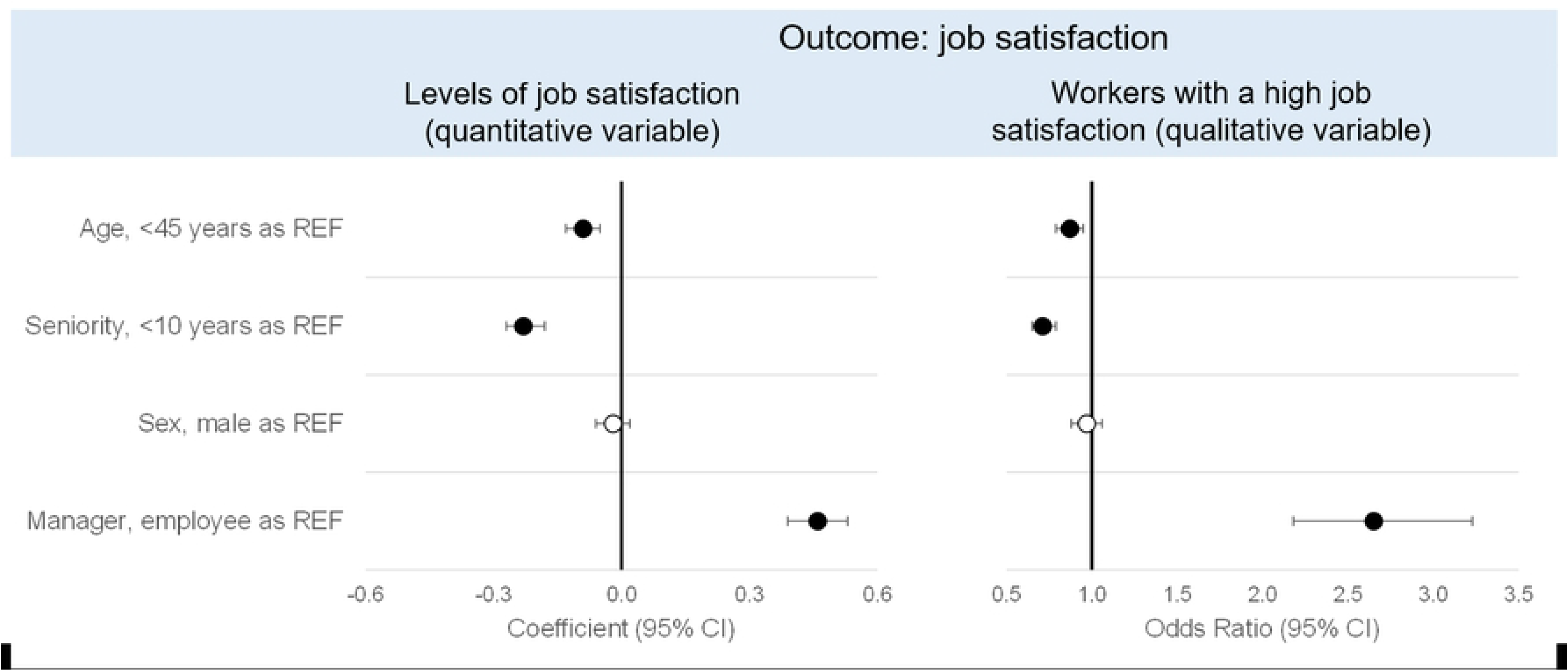
Influence of socio-demographic factors of job satisfaction A colored circle means that the measured effect is significant, an empty circle means no effect. The reference classes (REF) of the different parameters are the following: age: workers under 45; seniority: workers with less than 10 years of service; sex: male; job position: employee.

### Results of the “job satisfaction” questionnaire

Overall, the psychosocial feelings were higher in managers than in employees (**Fig 4**). Ambiance (mean ± SD 71.2 ± 20.9 vs 66.1 ± 24.2), meaning (66.9 ± 21.8 vs 56.1 ± 23.1), organization (55.3 ± 23.6 vs 46.6 ± 24.6), recognition (62.8 ± 23.9 vs 48.3 ± 26.7) and values (66.3 ± 21.7 vs 56.5 ± 23.1) were higher among managers (p < .001). Only the work-life balance (balance) did not seem to be impacted by the position within the company (58.1 ± 23.9 vs 59.2 ± 23.4, p = 0.2).

**Fig 4.**
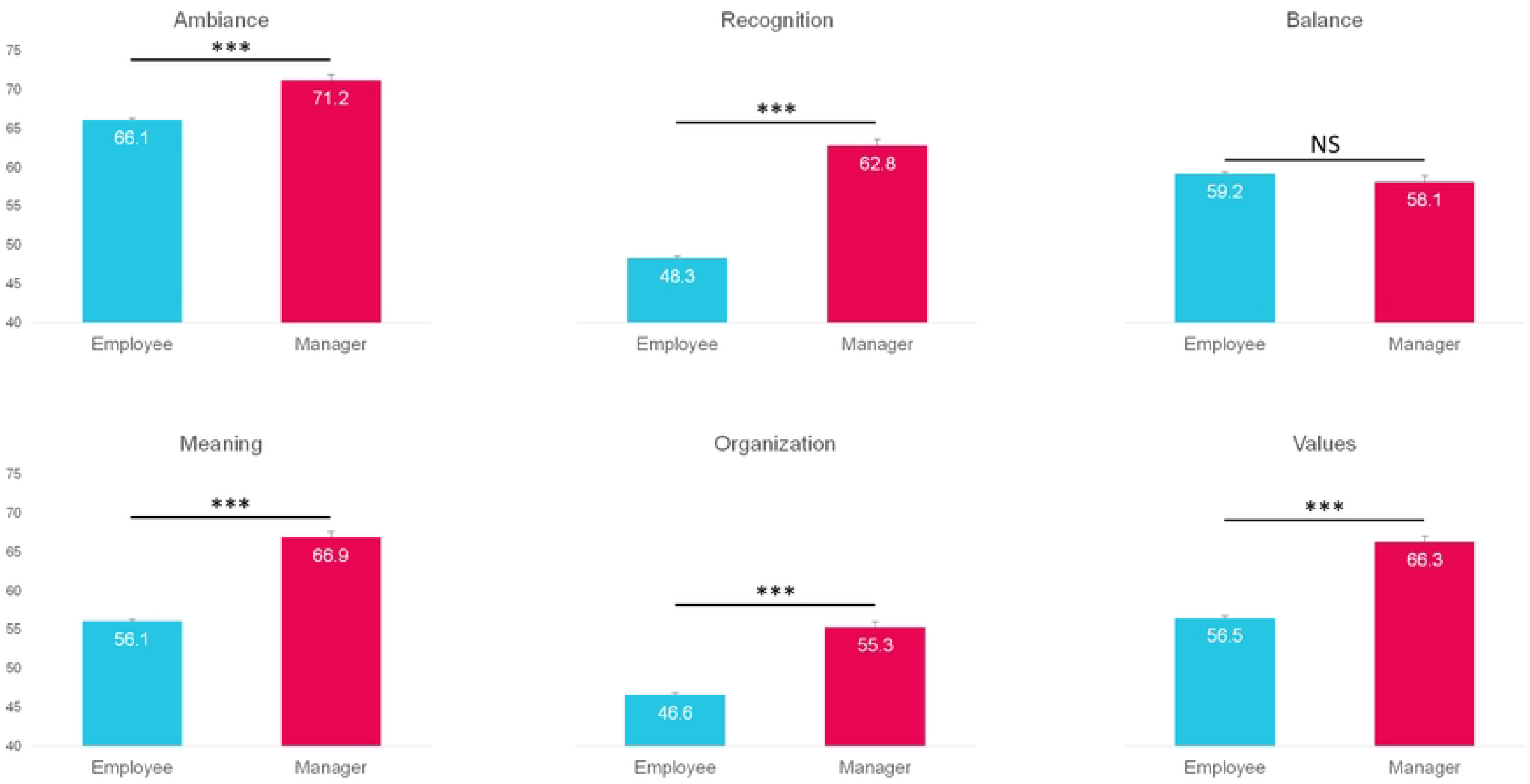
Psychosocial feelings by job position ‘***’: p < 0.001; ‘NS’: not significant

Each psychosocial factor was found to have positively influenced job satisfaction regardless of the workers job position (**Fig 5**). For managers, job satisfaction was mainly driven by recognition (ES = 0.48, 95% CI 0.33 to 0.58) followed by values (0.43, 0.30 to 0.57) and organization (0.36, 0.23 to 0.49) (p < .001). Among employees, recognition (0.45, 0.41 to 0.48), meaning (0.41, 0.37 to 0.45) and ambiance (0.32, 0.29 to 0.36) were the psychosocial factors that most influenced level of job satisfaction (p < .001). The balance was found to have the least contribution for both managers (0.18, 0.07 to 0.29) and employees (0.11, 0.07 to 0.14) (p < .001).

**Fig 5.**
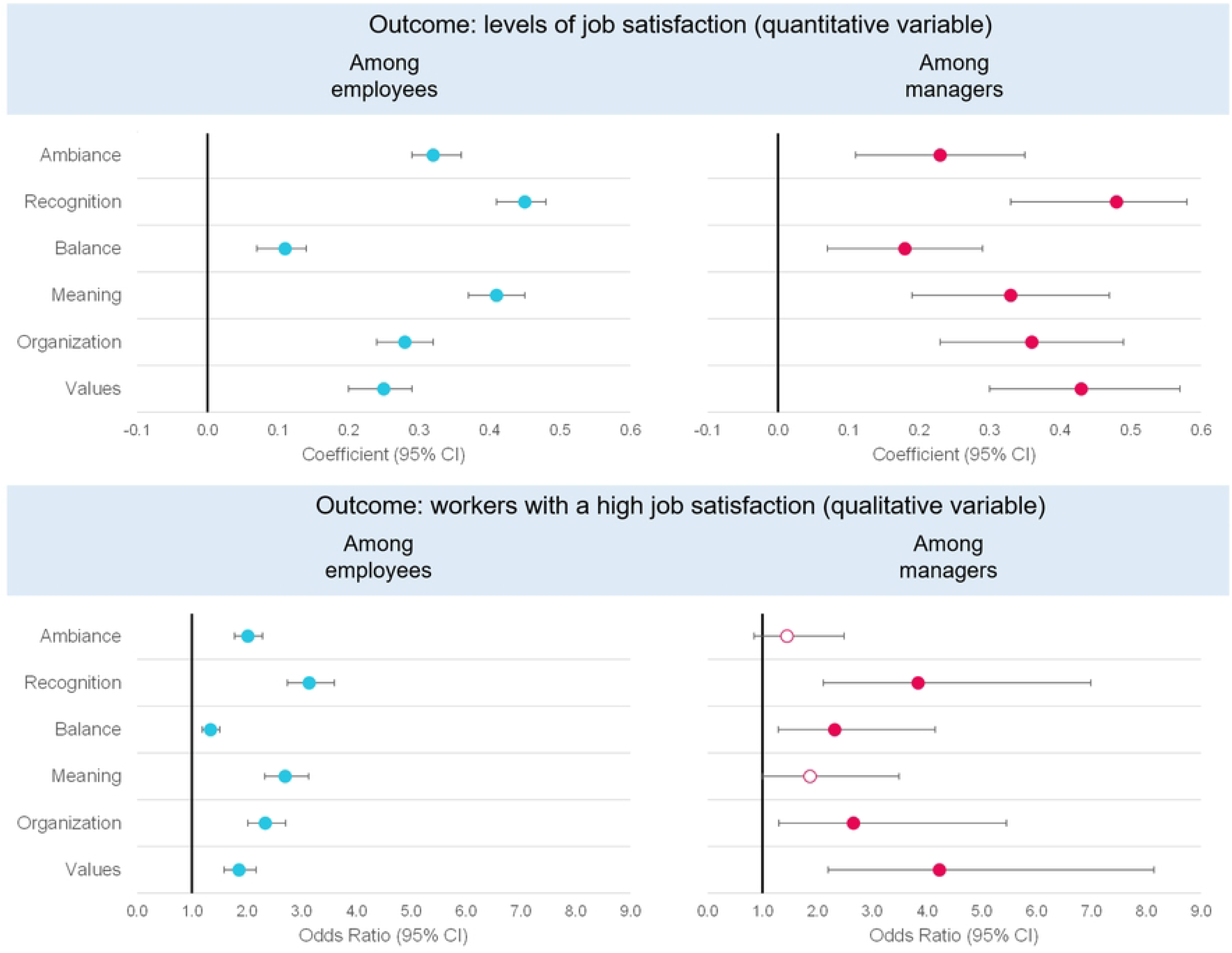
Influence of psychosocial factors of job satisfaction by job position A colored circle means that the measured effect is significant, an empty circle means no effect.

The job satisfaction indicators of recognition (OR = 3.84, 95% CI 2.11 to 6.99, p < .001), values (2.23, 2.20 to 8.14, p < .001) and organization (2.66, 1.30 to 5.45, p = 0.008) remained the primary drivers of a high job satisfaction in managers. Balance (2.32, 1.29 to 4.15, p = 0.005) and meaning (1.87, 1.00 to 3.49, p = 0.049) were also shown to be a driver of a high job satisfaction (2.05, 1.11 to 3.79, p = 0.02), but not the ambiance (1.45, 0.85 to 2.49, p = 0.2). In employees, recognition (3.14, 2.74 to 3.60) and meaning (2.70, 2.33 to 3.13) were the primary drivers of a high job satisfaction, followed by organization (2.34, 2.02 to 2.71) (p < .001). Other psychosocial factors, namely ambiance (2.02, 1.78 to 2.29), values (1.86, 1.59 to 2.17) and balance (1.34, 1.19 to 1.51) were also found to be drivers of a high job satisfaction in employees (p < .001) (**Fig 5**).

### The job satisfaction and the socio-demographic characteristics of the workers

In addition to the major effect of the managerial position, some socio-demographic variables have been revealed as factors of job satisfaction, although their effect was much smaller than that of the job position. Workers over 45 years of age (ES = -0.09, 95% CI -0.13 to -0.05) and those with more than 10 years of seniority (−0.23, -0.27 to -0.18) reported having a lower level of job satisfaction (p < .001) (**Figure 3**). Being over 45 years old decreases the chances of being satisfied at work by 13% (OR = 0.87, 95% CI 0.79 to 0.95, p = 0.002) and having 10 or more years of seniority by 19% (0.71, 0.65 to 0.79, p < .001). Finally, both job satisfaction level (−0.02, -0.06 to 0.02, p = 0.97) and prevalence of workers with a high job satisfaction (0.97, 0.88 to 1.06, p = 0.5) did not appear to differ by sex (**Fig 3**).

## Discussion

The main results show that in terms of job satisfaction and psychosocial feelings, the position within the company plays a crucial role. Psychosocial factors can influence the job satisfaction of managers and employees differently. Other socio-demographic factors were also found to have an impact on job satisfaction among workers.

### The influence of the manager’s role on job satisfaction

The role of the manager is crucial in the professional environment. Indeed, the manager has a heavy responsibility to achieve successful outcomes while ensuring that the expectations of their staff are met, both human and organizational (31,32). Therefore, the benefits of good leadership are numerous (3,33,34). Thus, if the role of the manager is to ensure the well-being and job satisfaction of their employees (35), it is also essential to ensure that they are satisfied themselves. In this study, we aimed to assess the influence of the managerial position on both job satisfaction and psychosocial feelings. Our results show that on average, managers have higher levels of job satisfaction and are more satisfied with their job than employees. Previously, differences in perceived job satisfaction according to position in a health care environment have shown that physicians had the highest job satisfaction (14), with registered nurses the lowest (36) among several healthcare professions. This may suggest that greater responsibilities attributed to managers, often associated with greater latitude in decision making, make them more satisfied than employees, who are themselves often the subject of management policies (37,38). Knowing that job satisfaction is linked to health benefits (1,2,4,5), organizations should strive to improve the job satisfaction of employees, who appear to be less satisfied than managers.

### Psychosocial feelings as factors of job satisfaction

All considered psychosocial factors were found to influence job satisfaction for both employees and managers. Our findings show that job satisfaction is positively influenced by ambiance at work, which is in line with literature that found that it can be improved by encouraging social support and team cohesion (36). Recognition at work can take several forms including notably recognition by patients or clients, by colleagues or by management. A study among doctors in China found that doctors with higher patient recognition presented higher level of job satisfaction (39), as observed in our study, with managers / employees reporting higher recognition having greater job satisfaction. Work-life balance was also found to be a factor of job satisfaction, which is in line with literature (40,41). A study conducted in health professionals found that work-family conflict can also lead to lower job satisfaction (42). Meaning at work is defined as “the discovery of existential meaning from work experience, work itself and work purpose/goals” (43). Thus, it is not surprising to find a direct link between meaning and job satisfaction as it translates among other things the motivation and the fulfillment of the worker. Organization commitment, known for being related to job satisfaction among nurses (13,44), was also found to be positively related to job satisfaction. Lastly, work values were positively related to job satisfaction, which is consistent with the findings of previous studies on the subject (45,46). It has also been highlighted among doctors that those with higher work values scores presented higher job satisfaction (39). Recommendations should be made for organizations to strive for improvement of workers’ psychosocial feelings, to consequently improve their job satisfaction.

### The socio-demographic factors of job satisfaction

Age and seniority also emerged as factors that could affect job satisfaction, with lower levels on average and even a lower prevalence of satisfied workers among the older workers (45 years old or more) and more senior workers (10 years of service of more). The link between age and job satisfaction has already been demonstrated, but the results are equivocal. Sometimes job satisfaction increased with age (47), sometimes it decreased (48) or presented a U-shape effect (49). For the seniority, its negative influence on job satisfaction has already been reported in the literature (47). No sex differences could be found in our study, although previous studies have shown that women pharmacists were more satisfied than men at work (50), while the opposite relationship has been found in physicians (51,52). Therefore, an absence of effect may not seem so surprising and suggests that, among Wittyfit’s users, the drivers of job satisfaction are the same for both men and women. Organizations must be aware that socio-demographic characteristics can influence job satisfaction to ensure that policies to improve well-being in the workplace are effective.

### Limitations

We acknowledge some limitations in our study. Self-reported feelings of workers (job satisfaction and psychosocial factors) were assessed using a single VAS instead of complete questionnaire as often used (53). This may lead to over- or under-estimated measurements that may cause a measurement bias as well as an affective bias (54). Nevertheless, with a Cronbach’s α of 0.84, we can assume that our questionnaire is reliable. Due to its size, our sample might not be representative of French workers. Indeed, it is not possible to state that it is representative in terms of jobs, nor even in terms of sectors of activity, as this information is not provided by Wittyfit’s clients. However, on the one hand, with the inclusion of seventeen different companies, it can be assumed that numerous jobs were represented. On the other hand, the inclusion of a random effect in our regression models, namely the “company” effect, allowed us to measure the influence of the company on the outcome independently of the fixed effects, namely the psychosocial and socio-demographic factors including the influence of the position held by the employee. Even taking these considerations into account, we believe generalizability of the findings from our study is possible.

## Conclusions

Managers have higher job satisfaction and psychosocial feelings at work than employees. Several psychosocial factors can influence workers’ job satisfaction. Associated effect sizes may vary according to the level of position within the company. Socio-demographic factors such as age and seniority may influence job satisfaction as well. Companies should implement policies aimed at improving the well-being of their employees to improve their job satisfaction, regardless of their position.

## Data Availability

Data from Wittyfit cannot be transmitted without the prior consent of the company’s corporate clients, except to the University Hospital of Clermont-Ferrand, France, which may use the data for research purposes.

## Acknowledgements

We express our sincere gratitude to all voluntary workers using Wittyfit, who participated in this study.

## Authors’ Contributions

Study design: FD, SD;

Study conduct: RC, FD, BP, SD;

Data collection: SD, TC;

Data analysis: RC, BP;

Data interpretation: RC, FD, BP;

Drafting manuscript: RC;

Revising manuscript content: RC, FD, BP, ACB;

Approving final version of manuscript: all authors.

RC, BP, SD, FD take responsibility for the integrity of the data analysis.

